# Show solidarity with the Congolese people in the 10th Ebola outbreak declared a health emergency of international concern: understand a qualitative study of variables of hospital activities on infection control practices in Kinshasa city

**DOI:** 10.1101/19000539

**Authors:** Guyguy Kabundi Tshima, Kaleb Tshimungu Kalala

## Abstract

Health workers play an important role during epidemics, but there is limited research on hospital activities on infection control practices in the Democratic Republic of the Congo and how health workers can cope during a probable health epidemic in Kinshasa city. The determinants of the current Ebola Virus Disease in the geographical distribution remain poorly understood. The World Health Organization’s Health Regulation Committee decided on Wednesday July 17^th^, 2019 to declare the Ebola haemorrhagic fever epidemic in the provinces of North Kivu and Ituri as a health emergency of international concern. The country struggles to control it against a backdrop of a health system that is already overburdened. To test the influence of the challenges of a contamination in the context of an Ebola outbreak that may face health workers and their coping strategies in thirteen hospitals of reference in Kinshasa, we conducted a survey hoping to educate or remember good practices for health workers in Kinshasa that is also available for health workers in the East Area of the country in which the ongoing Ebola outbreak progress is spreading (North Kivu and Ituri). For the ongoing outbreak, we obtained data from the Ministère de la Santé Publique of the Democratic Republic of the Congo where cases are classified as suspected, probable, or confirmed using national case definitions. **We found** that the ongoing Ebola virus outbreak in the Democratic Republic of the Congo has similar epidemiological features to previous Ebola virus disease outbreak in Sierra Leone that was well described. For the qualitative study about the biosecurity in thirteen hospitals of reference in Kinshasa, **we found** that the Bondeko-Ngaliema Monkole group has occupied the first rank, while the group Kintambo-King Baudouin-Ndjili-Makala occupied the other end of the scale; the other health facilities occupied an intermediate position. Among the 7 hospitals which were placed at the top of this classification of biosecurity, 5 were massively subsidized by international NGO, which explains to a great extent their performances in one hand, another hand finding its explanation in the quality of their management. It is the case of Bondeko, Monkole, Kalembe-Lembe, St Joseph and Kingasani 2.

**Author summary:** The determinants of the transmission are poorly understood, but a growing body of evidence supports an important role of the lack of prevention in the dissemination of Ebola virus. The results of our study conducted in 13 hospitals of reference in Kinshasa suggest that the biosecurity measures—which were introduced in Kinshasa hospitals policies through prevention since Ebola outbreaks—have been respected by 75% and had 25% of parameters to be improved. Biosecurity is an important concept; it seems to be a vector for the prevention of *Ebola Virus Disease*. In addition, the lack of biosecurity observation may have a role in the contamination of *Ebola Virus Disease* in local populations found in invaded areas. This study provides knowledge into the preventive measures influencing Ebola *Virus Disease* populations, thereby determining in perspective a study on meat consumption of animals found dead in forests that will be a risk for human infection as the Democratic Republic of the Congo has many forests.

## Introduction

### Background

There are five identified subtypes of Ebola virus. The subtypes have been named after the location where they have been first detected. Three of the five subtypes have been associated with large Ebola haemorrhagic fever (EHF) outbreaks in Africa. Ebola–Zaire, Ebola–Sudan and Ebola–Bundibugyo. EHF is a febrile haemorrhagic illness which causes death in 25–90% of all cases [1].

The Zaire strain is the most fatal, with an overall case fatality ratio ranging from 69% to 88% [1].

Ebola virus disease is transmitted primarily through contact with the body fluids of symptomatic patients, most commonly in adults aged 17–44 years, with relative sparing of children younger than 16 years [1,2].

The Democratic Republic of the Congo has had nine recorded Ebola virus disease outbreaks since 1976. The remoteness of most affected communities and the involvement of 2 deaths in Uganda, a neighboring country connected to the risk zone make this outbreak the most complex and high risk ever experienced by the Democratic Republic of the Congo [3].

### The ongoing outbreak

The ongoing outbreak occurred in January, 2019, in the North-Kivu Province, in the East of the country, causing a total of 1,449 deaths with 1,355 confirmed. Most of the previous outbreaks have been confined to remote rural areas, with the exception of an outbreak in Kikwit, a town with a population of nearly 400□000 that resulted in 315 cases and 250 deaths [4].

The ongoing response to the current outbreak in North-Kivu Province includes use of traditional measures such as early identification, isolation and care of cases, contact tracing, safe and dignified burials, and culturally appropriate community mobilisation (including risk communication, community engagement, and social mobilisation). These traditional measures are being supplemented by use of the recombinant vesicular stomatitis virus–Zaire Ebola virus (rVSV-ZEBOV) vaccine, with vaccination of first-line and second-line contacts [5].

### Specific objectives

This deep survey was conducted to inquire into what is concretely done regarding BS in the reference hospitals of the city of Kinshasa.

### Prespecified hypotheses

1. Transmission can be stopped by early diagnosis, patient isolation and care, infection control, safe and dignified burial of people who had died from Ebola virus disease, rigorous tracing of contacts, and, more recently, promising evidence on the use of targeted vaccination [2].
2. Being an ongoing-conflict country, the health system in the Democratic Republic of the Congo can be described as fragile and sub-optimal in the face of a disease outbreak as shown by its poor health outcomes [1].
3. In hospital settings, the primary level of infectious risk concerns the family members [6]. These naturally maintain diverse or intimate relationships with other members of their families thus exposing them to the secondary infectious risk. These in turn may transmit the infection to others in their neighborhood (tertiary infectious risk), and so on [7]. It brings the concept of biosecurity (BS).

### The concept of biosecurity (BS)

It brings together knowledge, attitudes and individual or collective practices in the rational management of biological, infectious or toxic risk. BS touches on many fields of activity, including the agro-food industries, animal production, air conditioning, drinking water supply, medical activities (care and vaccination), biomedical research, military… [8].

The common point is the risk of contamination by pathogenic microorganisms or biological products harmful to exposed individuals. The emergence of new pathogens as Ebola virus and the prodigious development of exchanges between countries and continents because of air transport raise the issue of BS in apocalyptic terms [9].

### Depth survey

In deep, BS concept incorporates a dozen variables, namely: (1) the respect of the standards of hygiene, (2) the posted written instructions, (3) the measures of individual protections (mask, glasses, gloves), (4) the cleanliness of hygienic facilities, (5) respect for the principles of teamwork, (6) asepsis, (7) sanitation, (8) basic education, (9) training, and (10) ease of access to care [10].

## Methods

This study was conducted between September and December 2003 and the last three months of March 2004 as a preliminary study. It used qualitative research methods - in depth interviews (IDI) with health workers and key informant interviews (KII) [2] – to explore workers experiences during their normal working days that is from 2004 to June 2019 [2]. Qualitative interviews facilitate generation of in-depth and contextual information about an individual’s experience, beliefs and perceptions as well as exploration of reasons behind their answers through probing questions [2].

The study was conducted in thirteen health facilities in Kinshasa. The selected study health facilities were selected for being the reference hospitals health, as they represent different health centers of Kinshasa. This survey targeted the emergency departments and laboratories of the 13 reference institutions in the city of Kinshasa.

### In depth interviews with health workers

Contextual information about the ongoing health situation in the country is checked using the press team service of communication of the Ministry of Health of the Democratic Republic of the Congo [1] as well as exploration of other media sources reasons through probing updates [2]. In the preliminary study, the investigators’ job was to record the results of their observations, as well as the answers to the questions provided by the health workers found at the targeted places (laboratories, emergency rooms) following a pre-established questionnaire. Our database contained 20 variables (Table 2).

**Table 1.** The epidemiological situation of the Ebola Virus Disease dated July 16, 2019.

**Table 2.** Exploration of variables of hospital activities on infection control practices.

### Key informant interviews

Doctors themselves were often surprised when asked if their blood pressure monitors and stethoscopes were disinfected after each patient consulted or if they washed their hands with soap or bleach before moving on to another patient.

### Data sources measurements

The interviews were conducted in French, in a private room in the health facility, office or in corridor where the participant felt most comfortable. Separate topic guides were used for the in-depth interviews with health workers and key informant interviews.

The data were synthesized in the form of principal components (PC) from which a projection map of the various hospitals following their BS performance was constructed. Each health facility was represented by one point. The proximity of the points on the map signified their resemblance, while their distance emphasized their dissimilarity. Hospitals were then grouped into a small number of categories given their similarity. This grouping was done using the hierarchical ascending classification technique described by Ward where the results appear as a dendrogram. The analyzes were performed using SPSS software version 10.0.7.

#### Efforts to address potential sources of bias

The data presented are subject to change after extensive investigation and after redistribution of cases and deaths in their respective health areas.

### Ethics

Ethical approval was obtained from the University of Kinshasa Scientific and Ethics Committee and the Tropical Medicine Department Research Ethics Committee. Rigorous informed consent process was followed: all participants were given verbal and detailed written information about the nature and purpose of the research before taking part; participants were made aware of their right to decline to answer questions, and were assured that measures are in place to anonymize responses. All participants gave their consent.

#### Quantitative variables

All data were anonymized to protect patient privacy. Regarding accessing patient data and the indication whether all human subjects were adult or whether a parent or guardian of any child participant provided informed consent on their behalf, this statement was not applicable as we work with data that were available from the Ministère de la Santé to inform people on the ongoing outbreak.

## Results

### Descriptive data

The epidemiological situation of the Ebola Virus Disease dated 16 July 2019: Since the beginning of the epidemic, the cumulative number of cases is 2,522: 2,428 confirmed and 94 probable. In total, there were 1,698 deaths (1,604 confirmed and 94 probable) and 717 people cured. 374 suspected cases under investigation; 10 new confirmed cases, including 6 in Beni, 2 in Mabalako, 1 in Katwa and 1 in Mangurujipa; 10 new confirmed cases deaths: 5 community deaths, including 3 in Beni, 1 in Mabalako and 1 in Mangurujipa; 5 deaths at Ebola Treatment Centers including 4 in Beni and 1 in Katwa; 7 people cured out of Mabalako Ebola Treatment Center. The cumulative number of confirmed / probable cases among health workers is 136 (5% of all confirmed / probable cases), including 41 deaths.

The objective of this paper was to educate health workers by revisiting or reminding the evaluation of the level of biosecurity (BS) in the emergency rooms and laboratories of 13 hospitals of reference of the city of Kinshasa.

In this problem, health facilities have a very special dimension. Indeed, they constitute real bio-reactors that concentrate, cross and multiply, by a factor of several orders of magnitude, the dangerousness of many germs brought by the patients or that circulate there.

Thus, any stay, even for a very short period, in these eminently dangerous places, can be of great consequence for an imprudent visitor, that directly or indirectly. In addition, gaseous or liquid effluents, as well as solid wastes emanating from these places pose a serious threat to the populations living in its immediate surroundings

### Details of hospital participants

Of the 13 health facilities surveyed, 6 had no instructions on BS, 9 had bleach in the laboratory and only 3 had bleach in the emergency room. In 7 hospitals, we saw active laboratory technicians wearing gloves; 10 had automatic pipettes. According to the answers of the health workers, the laboratory equipment was disinfected the same day in 12 health facilities. Ten regularly emptied their baskets. Seven had an incinerator. Room ventilation was satisfactory in 9 cases and enough lighting in 12 cases (Table 2).

In emergency rooms, examination equipment (sphygmomanometer, stethoscope and thermometer) was not regularly disinfected after use in only 2 facilities. The change of sheets after examination of each patient was only observed in 4 establishments. In 2 cases, the examiners wore gloves. The toilet was well kept in 5 cases (Table 2).

Overall, for each parameter studied, the achievement performance for all health facilities was classified into 4 categories: excellent, if performed by more than 75% of hospitals; average, if carried out by 50 to 75% of hospitals; low, if carried out by 25 to 50% of hospitals; bad, if done by less than 25% of hospitals.

Thus, the following 6 parameters had obtained the excellent score: disposable sampling equipment; disinfection of equipment and the bench; good lighting of premises; waiting room unencumbered; adequate preservation of biological specimens; regular emptying of baskets.

In the extreme, the following 2 parameters were rated bad: gloves worn by examiners; disinfection of examination instruments.

The other parameters had a rating from low to medium.

#### Hospitals profile

The 1st principal component (PC) represented the direction of the increase of the BS performance at the top of which the Bondeko-Ngaliema-Monkole group was standing, as opposed to the other hospitals surveyed, notably the Makala-Ndjili-King Baudouin group. Kintambo in extreme position. The Kinshasa General Hospital was in a position close to the middle.

The 2nd PC represented the direction of the increased contrast between labs and emergency rooms in terms of BS.

Following this axis, Monkole, Kintambo, Bondeko, King Baudouin and Ngaliema were opposed to other hospitals.

Considering the groupings provided by the dendrogram, it was noted that for a relative distance of 15 units, health facilities formed the following three aggregates:

⍰ Group 1: Makala, King Baudouin, Ndjili and Kintambo

⍰ Group 2: Ngaliema, Bondeko

⍰ Group 3: General Hospital, Monkole, Kingasani, Kinoise, CUK, Kalembe-lembe, St Joseph

Group 1 was away from the other two groups by 19 units of distance. Thus, a relative distance of 10 units corresponded to the following 6 aggregates:

⍰ Group 1: Makala

⍰ Group 2: King Baudouin, Ndjili, Kintambo

⍰ Group 3: Ngaliema, Bondeko

⍰ Group 4: Kinshasa General Hospital

⍰ Group 5: Monkole, Kingasani, Kinoise

⍰ Group 6: CUK, Kalembe-lembe, St Joseph

Following this second grouping, it was noted that Makala stood out from the group it formed with King Baudouin and Kintambo.

Former group 3 was broken into three pieces (groups 4, 5 and 6). The General Hospital of Kinshasa was isolated from the others.

The group Monkole-Kingasani-Kinoise (group 5) was away from the group CUK-Kalembe-lembe-St Joseph (group 6) by 11 units of distance.

Considered according to the performance achieved, the group Bondeko-Ngaliema (group 2 in the first ranking) had the best score with 15 parameters of excellent level, 4 of average level and none of mediocre level.

In second place came Group 3 with 10 excellent level parameters and 3 parameters at each of the lower levels.

In third place was group 1, which had 5, 3, 4 and 7 parameters, respectively excellent, medium, low and mediocre.

The difference in the distribution of performance frequencies between groups was highly significant (chi-squared = 12.5, 2 degrees of freedom, p = 0.002).

## Discussion

### Key results

This deep survey was conducted to inquire into what is concretely done regarding BS in the reference hospitals of the city of Kinshasa [11]. It showed that health facilities in Kinshasa have a very variable level of BS ranging from excellent to poor [13].

The Bondeko-Ngaliema-Monkole group undoubtedly occupied the first rank, while the Kintambo-Roi Baudouin-Ndjili-Makala group occupied the other end of the scale; the other hospitals were in an intermediate position.

It is important to note that the University Hospital of Kinshasa is placed 8th in this classification, between the Kingasani Hospital and the General Hospital. It is far from the time when this hospital was presented as “the top of medicine” in the DRC. The consequences of such a fall are incalculable when one thinks that this hospital more than another is eminently involved in the process of training doctors in our country.

Its deliquescence is only the logical reflection of the difficulties known to all that our country is currently going through. Of the seven hospitals that placed before the University hospital in this classification of BS, five are heavily subsidized by international NGOs, which largely explains their performance, another party finding its explanation in the quality of their management. They are: Bondeko, Monkole, Kalembe-Lembe, St Joseph and Kingasani 2.

### Limitations of the study, taking into account sources of potential bias or imprecision. Discuss both direction and magnitude of any potential bias

To be contaminated, there is no need to transgress all the elements of this chain of security: the transgression of only one of them is enough to lead to a catastrophe. The wearing of gloves in the laboratory was highly rated with a rate of 83.8% for all health facilities. It is a manifestation of an awareness translated positively act against the risk of infection. This good practice has been adopted by the maintenance staff to the point of perverting it. Indeed, we even saw a cleaner, with gloved hands, move from one room to another opening doors after doors, without anyone having to call him.

The state of the toilets, often disastrous in many of the hospitals, is betrayed by the pestilential odors that emerge, and the presence of the bins not regularly emptied, besides the presence of the cockroaches, makes the stay in the hospital more that formidable.

## Key informant interviews

Doctors themselves were often surprised when asked if their blood pressure monitors and stethoscopes were disinfected after each patient consulted or if they washed their hands with soap or bleach before moving on to another patient.

## Data collection and analysis

The interviews, were conducted in French, in a private room in the health facility, office or in corridor where the participant felt most comfortable. Separate topic guides were used for the in-depth interviews with health workers and key informant interviews. The topic guides for the in-depth interviews covered health workers’ perceptions and experiences of working during their normal working day, any constraints that they faced, challenges in the health systems, their coping mechanisms, and options to increase the resilience of workers and the health system in the future. The topic guides for the key informants included the following areas: perceptions and experiences of the risk contamination, of Ebola outbreak; its impact on health workers; constraints, challenges and opportunities in relation to leadership and governance, health workforce and service delivery during the normal working day; and options to increase the resilience of workers and the health system in the fear of an Ebola epidemic phase.

### A cautious overall interpretation of results considering objectives, limitations, multiplicity of analyses, results from similar studies, and other relevant evidence

The interviews were manually recorded after gaining permission from the participants. The recordings of the interviews were transcribed verbatim and analysed using the framework approach which facilitates rigorous and transparent analysis. The coding framework was developed using themes emerging from the data, the topic guides and study objectives. The authors applied the coding framework to the transcripts, Ministry of Health DRC Press team update information was used to support the analysis.

### The generalisability (external validity) of the study results

In the great majority of laboratories, according to workers answers, disinfection of the bench would be done regularly, practically every day with alcohol. The disinfection of the instruments, the bench is not a simple question. It must be subject to a rigorous protocol specific to each case. The products used are chosen according to their effectiveness against microorganisms, their safety and their non-reactivity with the equipment. This aspect of the issue has not been addressed in this study.

The use of disposable sheets is also not a widespread practice in our hospitals, to the point where we have seen the patients’ parents spontaneously use their loincloths to have their sick child examined.

Other considerations include the cockroaches that are legion in hospitals: These undesirable companions of man deserve a whole study. In fact, these intrepid insects impose their law in the buildings they occupy, passing sewers at any place where they are not expected, making a real problem face to all BS measures taken in hospitals [14].

The promiscuity that reigns in our emergency rooms adds an extra dimension to the drama that is playing out. The patients, not isolated from each other, are seated on benches, for those who can, where most of the time they are on the floor, joined together, pell-mell, according to the chance of the arrivals. In some hospitals, an effort has been made to ensure that each patient has an individual plastic chair that is easy to clean and disinfect.

Those who did not have were simply examined by the leatherette or what was left of it. The risk of transmission of the pathogenic germs of a patient is, so to speak, amplified dramatically. We even surprised a head of Department to give instructions, during the visit of an authority of the place, so that the beds of the patients are covered with sheets clean and new leaves to remove them immediately after the visit.

Authentic! Some of our young doctors have never seen a clean sheet throughout their training. What do they know about BS if no one has ever talked to them about it, as this is not part of their curriculum? They will reproduce, where they will be, what they have been taught unfortunately carelessness. The responsibility of trainers and public authorities is total.

The lack of an incinerator is another observed aberration whose consequence on the environment is immeasurable. Who can understand that a hospital may lack an incinerator? And yet, it is unfortunately the case. This question is part of the problem of waste management at the health facility level. Liquid waste is discharged through public sewers; solids are either buried or simply thrown into dumps in the immediate vicinity.

In fact, no regulation seems to compel health facilities to manage orthodox waste. As if that were not enough, it is also necessary to add to this tragedy the almost chronic water supply disruption that strikes severely all the institutions that do not have enough water reserves. On this point, the excellence rating goes to the Monkole Hospital Center, which has an artesian well that protects it from untimely and prolonged breaks in water supply through the network to which it is connected.

It seemed to us that compliance with BS’s instructions was, so to speak, left to everyone’s discretion. We did not feel the existence of a constraint forcing the different actors to pay more attention. For example, while almost all facilities have internal or external refectories, in some hospitals it was quite amazing to see caregivers or health workers take their meals on the job site itself.

What can be the life expectancy of health workers in such conditions? The absence of instructions posted does not contribute to the promotion of the BS. Having instructions posted is one thing, respecting them is another.

This is a deep survey, however, highlighted serious failure to meet BS standards in most referral hospitals. This necessitates the taking of urgent decisions at all levels of responsibility so that eventually a BS audit is instituted within each hospital. The latter will be planning a more in-depth exploration of impacts of hospital activities on infection control practices [12-23].

There are serious deficiencies in contamination risk communication and infection in the Democratic Republic of the Congo. For example, the meaning of “Draconian precautions should be taken in the health facilities to protect all people called to evolve participate in the concept of biosecurity (BS)” is unknown maybe it is an expression appropriate for a scientific report. Overall, it is interest to publish such paper with the appreciation of the relevance of the biosecurity findings to the Ebola outbreak [24,25].

### Meeting of the Emergency Committee of the International Health Regulations

The meeting of the Emergency Committee for International Health Regulations, convened by the Director-General of WHO, was held on Wednesday, July 16, 2019. The Emergency Committee concluded that the Ebola outbreak in the provinces of North Kivu and Ituri constitute a public health emergency of international concern At the meeting of the committee, the Director General of WHO, Tedros Adhanom, asked the international community to show solidarity with the Congolese people. He also asked not to impose “punitive and counterproductive restrictions that will only serve to isolate the DRC at the international level”. “These restrictions on travel and trade will be useless. There have already been more than 75 million people controlled at the border posts,” he said at a press conference, sanctioning the meeting [26].

The Ministry of Health accepts the evaluation of the expert committee. The ministry hopes that this decision is not the result of the many pressures from different stakeholder groups who wanted to use this statement as an opportunity to raise funds for humanitarian actors despite the potentially harmful and unforeseen consequences for the affected communities that depend on them greatly from cross-border trade for their survival [26].

Furthermore, we regret that after spending almost a year in this epidemic, certain groups of people in the community continue to adopt irresponsible behavior that causes the geographical spread of the virus. It is important to remember that in the cases of Goma and Uganda, the patients knew that they were at risk but refused to respect the health recommendations and deliberately traveled to another area. The Government will consider what steps need to be taken to prevent these high-risk groups from continuing to spread the epidemic in the region [26].

### Follow-up of the situation of the pastor’s contacts who traveled to Goma

Vaccination around the confirmed Goma case continues at the Afia Himbi Health Center in the Goma Health Zone. All contacts in the city were found in less than 72 hours, including the motorcycle taxi driver that the pastor had used to get to the health center. The response teams from Beni and Butembo continue the investigations to trace the pastor’s journey and identify his contacts in these two cities [26].

The Committee noted that epidemiological trends are evolving positively in the Butembo and Katwa epicentres. However, the response is beginning to face challenges related to community acceptance and security in areas such as Mabalako where the epidemic has resumed recently. On the other hand, the response continues to be limited by the lack of adequate funding and limited human resources [27].

The Committee commended the communication and collaboration between the DRC and Uganda that rapidly contained Ebola cases that crossed the border. Uganda’s rapid response proves the importance of preparing border countries for DRC [26].

After lengthy discussions, the Committee considered that the criteria for declaring a public health emergency of international concern were not fulfilled. The Committee considered that all public health measures and recommendations needed to end the epidemic were already being implemented by WHO and the affected countries [26].

## Conclusions

We address an important topic: contamination in the context of an Ebola outbreak. We offer a survey of infection control practices and suggest more in-depth exploration of the impacts of hospital activities on infection control. There are serious deficiencies in contamination risk communication and infection in the Democratic Republic of the Congo. Overall, there is interest in publishing a such paper with the appreciation of the relevance of the biosecurity findings to the Ebola outbreak. Our investigation did not address or deepen all aspects of BS. We did not investigate all hospital sectors (delivery rooms, internal medicine, pediatrics, surgery, etc.).

## Perspective

### The Social responsibility in the control of an infection risk

While the Government continues to openly share with partners and donors the way in which it uses the funds received, we hope that there will be greater transparency and accountability of humanitarian actors in their use of funds to respond to this Ebola outbreak

The Ebola epidemic is above all a public health crisis that requires a response by actors with real technical expertise. However, the main difficulty is that this epidemic occurs in an environment characterized by problems of development and shortcomings of the health system.

### Communication on risks contamination in meat consumption of dead animals

We hope to conduct a KAP (Knowledge, Attitude and Practices) survey of health workers, including students, on biosecurity issues, particularly communication on risks contamination in meat consumption of animals found dead in forests, and issues related to the transmission of Ebola Virus disease and food insecurity.

The overall aim for this perspective will to develop the foundation of an education campaign on prevention of risks and the methodology for a comprehensive study of the communication risks and the evaluation of community behaviour change in the Democratic Republic of the Congo about eating meat from animals found dead in the forest because the Democratic Republic of the Congo has many forests. We will analyse that rising incidence and large-scale outbreaks are largely due to foods insecurity, inadequate living conditions, naïve populations, meat trade and population mobility, climate change making the hunters, the principal vectors of contaminated meat of dead animals.

Thinking on meat risk contamination was evidenced by the past Ebola Virus Disease outbreaks. We will highlight the first goal of sustainable development: no poverty because based on poverty, the interdiction of eating meats of dead animals (in forest which may be identified responsible for transmitting Ebola Virus Disease) cannot work as these circulating contaminated meats are difficult to control without means and efficiently transmit diseases.

## Data Availability

The dataset is available in the manuscript in tables. Add information on the ongoing Ebola outbreak is available from the corresponding author on reasonable request.

## Acknowledgements

We thank Drs: Zinszer Kate, Mvumbi Lelo Georges, Mafuta Eric, Ahuka-Mundeke Steve, Justus Nsio, Kalala Lunganza Richard, Odio Wobin Taddhée, Bobanga Lengu Thierry, Situakibanza Nani-Tuma Hippolyte, Lutumba Pascal, Kayembe Ntumba JM, Mulumba Madishala Paul, Kisasa Kafutshi Robert, Sylvie-Mireille Bambi, Onyamboko Akatshi Marie, Lebwaze Massamba Bienvenu, Kamangu Ntambwe Erick, Wumba Roger, Mitashi Patrick, Kayembe Kalambay Patrick and Mr Mukinay Tumb’ Tumb’ Nestor.

## Availability of data and materials

The dataset is available in the manuscript in 2 tables. Add information on the ongoing Ebola outbreak is available from the update message of Press Team of the Ministry of Health Democratic Republic of the Congo on reasonable request.

## Abbreviations

EVD: Ebola Virus Disease
CTE: Centre Traitement Ebola
HW: Health worker
IDI: In-depth interview
PC: Principal Component
KI: Key informant
KII: Key informant interview
BS: Biosecurity

## Authors’ contributions

GTK contributed to the design of the study, coordinated the data collection, supported the analysis and interpretation of the data and drafted the manuscript. KTK contributed to the design of the study, the analysis and interpretation of the data, and reviewed update drafts of the manuscript. All authors read and approved the final manuscript.

## Notes

### Consent for publication

Not applicable.

### Contributor Information

Guyguy Kabundi Tshima, (institutional); (private)

Kaleb Tshimungu Kalala

S1 Checklist: STROBE Checklist

